# Health service inequalities during the COVID-19 pandemic among elderly people living in large urban and non-urban areas in Florida, US

**DOI:** 10.1101/2020.05.01.20087791

**Authors:** Xinhua Yu

## Abstract

**Objectives:** Health inequalities were often exacerbated during the emerging epidemic. This study examined urban and non-urban inequalities in health services among COVID-19 patients aged 65 or above in US Florida from March 2 to May 27, 2020.

**Methods:** A retrospective time series analysis was conducted using individual patient records. Multivariable Poisson and logistic models were used to calculate adjusted incidence of COVID-19 and the associated rates of emergency department (ED) visits, hospitalizations and deaths.

**Results:** As of May 27, 2020, there were 13,659 elderly COVID-19 patients (people aged 65 or above) in Florida and 14.9% of them died. Elderly people living in small metropolitan areas might be less likely to be confirmed with COVID-19 infection than those living in large metropolitan areas. The ED visit and hospitalization rates decreased significantly across metropolitan statuses for both men and women. Those patients living in small metropolitan or rural areas were less likely to be hospitalized than those living in large metropolitan areas (35% and 34% versus 41%). Elderly women aged 75 or above living in rural areas had 113% higher adjusted incidence of COVID-19 than those living in large metropolitan areas, and the rates of hospitalizations were lower compared with those counterparts living in large metropolitan areas (29% versus 46%; OR: 0.37 [0.25-0.54]; p <0.001).

**Conclusions:** For elderly people living in US Florida, those who living in small metropolitan or rural areas were less likely to receive adequate health care than those who living in large or medium metropolitan areas during the COVID-19 pandemic.

## Introduction

Since December 2019, the novel Severe Acute Respiratory Syndrome associated coronavirus (SARS CoV2) (Zhu et al. 2020) has infected over 6 million people and claimed more than 370,000 lives worldwide (JHU 2020). Unlike the 2003 SARS virus that had limited transmissibility before symptom onset (Peiris et al. 2003), the novel SARS CoV2 can be transmitted from pre-symptomatic and asymptomatic patients (Bai et al. 2020; Huang et al. 2020b; Li et al. 2020) and cause sudden symptom exacerbation among mildly symptomatic patients, often leading to cytokine storm and acute respiratory distress syndrome (ARDS) (Guan et al. 2020; Huang et al. 2020a). The unprecedented scale of pandemic has forced many countries to adopt aggressive mitigating measures such as social distancing, closing schools and business, and prohibiting large gatherings (Anderson et al. 2020; Ferguson et al. 2020; Pan et al. 2020). Consequently, the epidemic in the US has slowed down significantly and many metropolitan areas have reached a turning point with reproduction numbers of one or below after April 15, 2020, as demonstrated in our previous study (Yu 2020).

People aged 65 or above were disproportionally affected by the pandemic, as about 80% of deaths occurred among this group (referred as elderly people in this report) (Garg et al. 2020). Due to their physiologically weak immunity and high prevalence of comorbidities in which two thirds of elderly people had two or more chronic conditions (Chavan et al. 2017), elderly people might be more likely to have severe disease if infected by the virus. In addition, timely diagnosis was critical during the COVID-19 pandemic, as early diagnosis and treatment might allow early interventions to reduce the risk of developing ARDS. However, timely diagnosis and treatment might be impeded by myriads of health care access barriers such as lack of transportation, difficulties in communicating with health care providers, and the complexity of the health care system (Fitzpatrick et al. 2004; Hill et al. 2015). For example, in a Netherlands study, elderly breast cancer patients were more likely to diagnosed with high stage of cancer and less likely to receive surgical treatment (Bastiaannet et al. 2010). During the COVID-19 pandemic, many elderly patients living in New York, US had prolonged stays in the intensive care unit and one fourth of them eventually died (Richardson et al. 2020).

Unfortunately, as history has shown, health inequalities might be exacerbated during the emerging epidemic (Hill et al. 2015). Awakened by this, many states started reporting the numbers of cases, hospitalizations and deaths by age and racial/ethnicity groups, revealing a disproportionally heavier disease burden among vulnerable elderly populations and among African Americans and other minority groups (FL-DOH 2020a; Santich 2020). Health inequalities could be due to differences in socio-economic status (SES). People with lower SES were more likely to have two or more chronic conditions than those with higher SES, (Kail et al. 2020; Singer et al. 2019) and may have limited access to health care resources. Therefore, SES factors could play an important role in survival among elderly people during the COVID-19 pandemic.

One particular aspect of health service inequalities is related to the differences between urban and non-urban areas. Not only are health care resources less sufficient in non-urban areas than urban areas, but also socioeconomic status and age distributions are different between them (de Boer et al. 2019). Elderly living in non-urban areas were more likely to receive inadequate health care than elderly living in large urban areas (Martino et al. 2019), and elderly colorectal cancer patients living rural areas were on average 18 days longer to receive a diagnosis than elderly living in urban areas (Bergin et al. 2018). Meanwhile, despite wide availability of maps representing the epidemic process, little was known about how geographic differences affected the health inequalities among elderly people living in large, medium, small metropolitan or rural areas. Existing reports and maps often focused on the description of the epidemic (e.g., websites driven by a GIS system like that of Johns Hopkins University (JHU 2020)), but none has carefully explored the inequalities underlying the reported case counts with appropriate epidemiological methods.

In this study, we aimed to examine urban and non-urban inequalities in health services during the COVID-19 pandemic among elderly patients in US Florida from March 2 to May 27, 2020. Since urban areas have more health care resources than non-urban areas, and those living in non-urban areas are more likely to have lower SES, we hypothesized that those living in small metropolitan or rural areas might have lower rates of visiting an ED, being hospitalized and higher mortality rates than those living in large or medium metropolitan areas.

## Methods

### Data sources

All lab confirmed COVID-19 cases were listed online by the Florida Department of Health (FL-DOH 2020a) and pre-processed (Aden-Buie 2020). The line list file included patient’s county, age, gender, residency, case confirmation date, contact history, ever visited an ED, being hospitalized and death status. However, no dates for ED visits, hospitalizations, or deaths were explicitly recorded in the file.

Metropolitan status for each county was obtained from National Center for Health Statistics (CDC 2020). Florida population data for 2018 were obtained from Florida health charts website (FL-DOH 2020b). We merged all these data sources by gender, age groups, and county.

### Statistical analysis

This is time series of COVID-19 cases in Florida. The first case was recorded on March 2, 2020. As of May 27,2020, there were 53,285 confirmed COVID-19 cases. We excluded 23 patients who did not have age information, and additional 86 patients without county information, resulting in 53,176 COVID-19 cases of all age groups and 13,659 cases aged 65 or above (referred as elderly people in this report) included in the analysis.

Patient’s age was grouped into <25, 25-44, 45-64, 65-74, and 75+. However, except for Table 1 and Figure 1 which presented an overview of COVID-19 epidemic in Florida, our main analyses were restricted to patients aged 65 or older, as the focus of this study was about health disparities among elderly people.

**Table 1:**
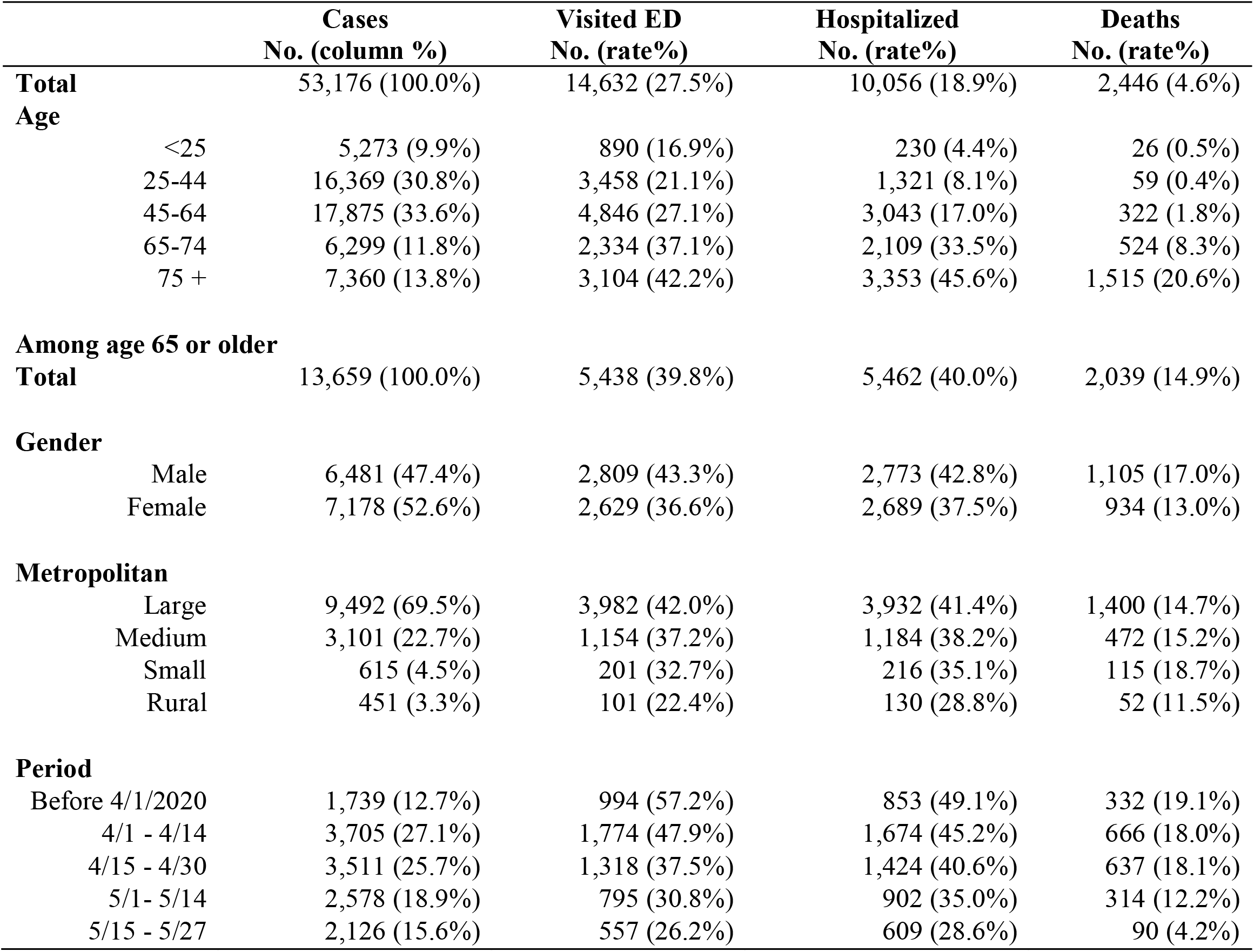
Descriptive characteristics of COVID-19 cases diagnosed in Florida as of May 27, 2020

**Figure 1:**
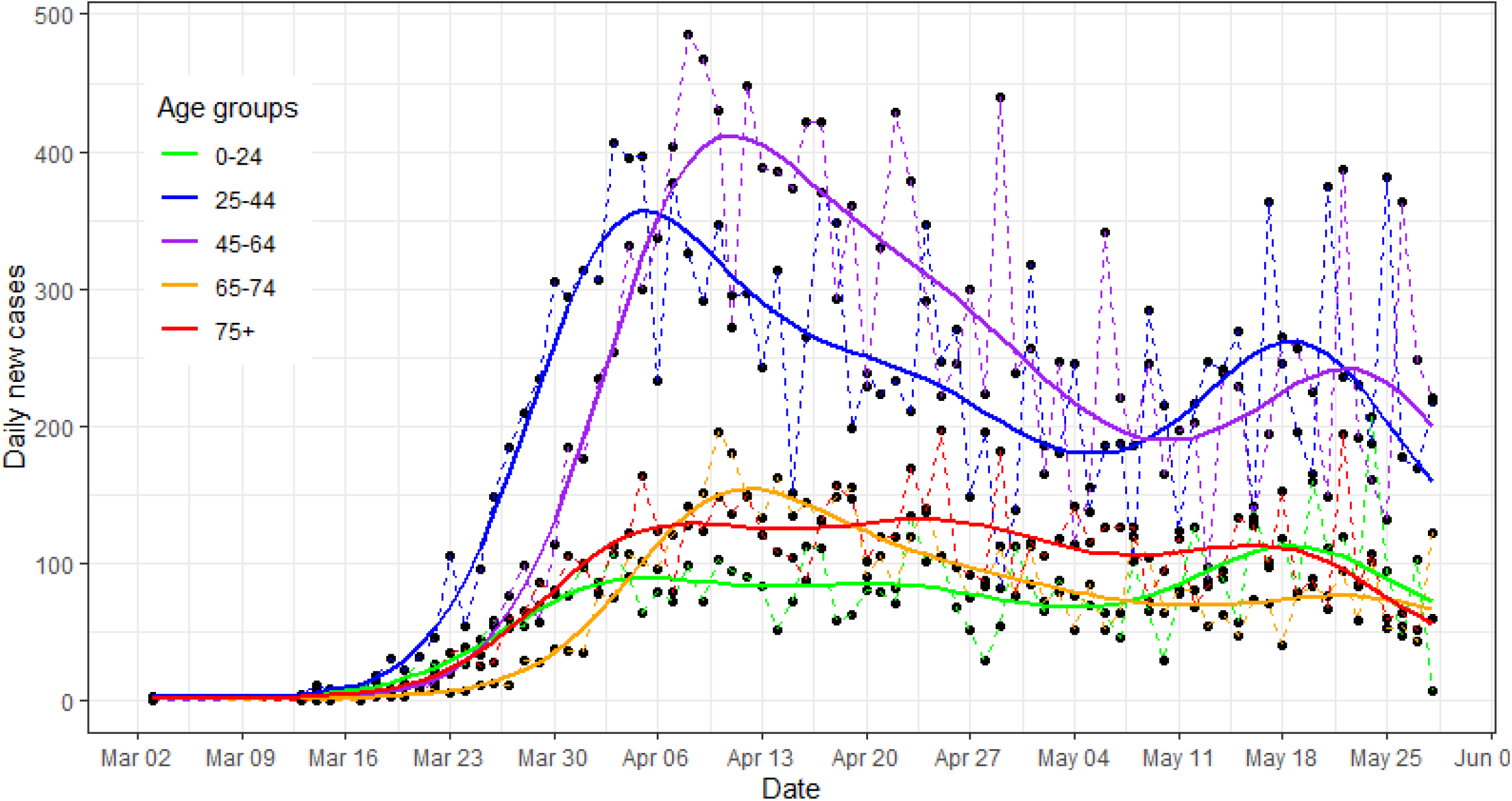
Fitted COVID-19 epidemic curves by age groups in Florida, as of May 27, 2020

Metropolitan status of each county was classified as large metropolitan areas and their suburbs (1 million or more people), medium metropolitan areas (250,000 – 1 million), small metropolitan areas (50,000 – 250,000), and non-metro areas (counties with 50,000 or less people, broadly considered as rural areas in this report). The number of cases were listed in the appendix table by metropolitan status (Appendix Table 3).

We also divided the whole epidemic into five periods: before April 1, 2020 and every two weeks thereafter (Table 1). After April 1, 2020, many control measures were enforced, including stay-at-home rule issued by the Florida state government on April 3, 2020. All models were adjusted for periods.

In addition to descriptive statistics about the number of cases, COVID-19 associated ED visits, hospitalizations and deaths, we calculated adjusted incident rates (per 1,000 persons) for COVID-19 based on Poisson regressions. The independent variable in the Poisson regressions was case counts by age and gender groups with age and gender specific population in each county as the proper denominator. Therefore, the predicted rates from the above Poisson regressions were the adjusted incidence by adjusting for age and pooling over all counties. In addition, using the line list file for individual COVID-19 cases, we employed logistic regressions to calculate probabilities (rates) of ED visits, hospitalizations, and deaths. The independent variable of the logistic regressions was the status (0/1) of ED visit, hospitalization or death for each COVID-19 patient, adjusting for age, gender, period and county. The adjusted rates were obtained from the predictive margins of the models. Furthermore, we predicted age specific epidemic curves from semi-parametric generalized additive models with smoothed time terms, assuming daily new cases follow a negative binomial distribution. We also mapped the adjusted hospitalization rates (per 100 cases) for each county based on a Poisson model.

SAS 9.4 (SAS Inc, Cary, NC), Stata 16.1 (Stata Inc, College Station, TX) and R *mgcv* package (Wood 2017) were used in the analysis. Although we set the large metropolitan area as the reference group for most of our comparisons, Bonferroni adjustment was also used to account for multiple comparisons, resulting a significance level of p < 0.01 for all comparisons.

Data and codes are available online (https://github.com/xinhuavu/urbanruralFL)

## Results

Table 1 presented an overview of COVID-19 epidemic in Florida as of May 27, 2020. Of 53,176 confirmed cases, 27.5% of cases had visited ED, 18.9% were hospitalized, and 4.6% died. Majority of cases were in metropolitan counties (Appendix Table 3). Although cases aged 65 or older accounted for only 25.6% of total cases, they accounted for 54.3% of hospitalizations and 83.4% of deaths. They were two to three times more likely to be hospitalized, and five to ten times more likely to die than people aged 45-64 (Table 1).

Among those 13,659 elderly patients, about 40% of them visited an ED and 40% of them were hospitalized. About 14.9% of them died (Table 1). Elderly men were slightly more likely to visit an ED, be hospitalized and die than elderly women. About 7.8% of elderly patients lived in small metropolitan or rural areas. They had lower unadjusted rates of ED visits and hospitalizations compared with those living in large or medium metropolitan areas. Those who were diagnosed before April 1 were more likely to visit an ED, be hospitalized, or die than those who were diagnosed after April 1, possibly because more mildly symptomatic patients were detected in late periods. The lower rates of ED visits, hospitalizations and deaths during the last period (after May 15) were more likely due to reporting delays.

Figure 1 presented the epidemic curves by age groups. The epidemic among people aged 25-44 seemed to lead the epidemic in the whole population, followed by those aged 45-64 and those aged 65-74. These groups had experienced two peaks: one major peak around April 5-10, and a small one around May 15-20. However, the daily new cases among people aged 75 or above remained stable from around April 1 to around May 15. Similar patterns existed in the epidemic curves by metropolitan statuses among elderly people (Appendix Figure 1).

The adjusted incident rates of COVID-19 and adjusted rates of ED visits, hospitalizations and deaths among COVID-19 patients were presented in Figure 2a-d (details in Appendix Table 1 and Table 2). Overall, the differences in adjusted incident rates between elderly men and women were small, while elderly men were more likely to have an ED visit (p<0.01 for men vs. women aged 75 or above), be hospitalized or die with COVID-19 than elderly women of the same age group and living in the same areas, though many comparisons were not statistically significant after Bonferroni adjustment.

**Figure 2:**
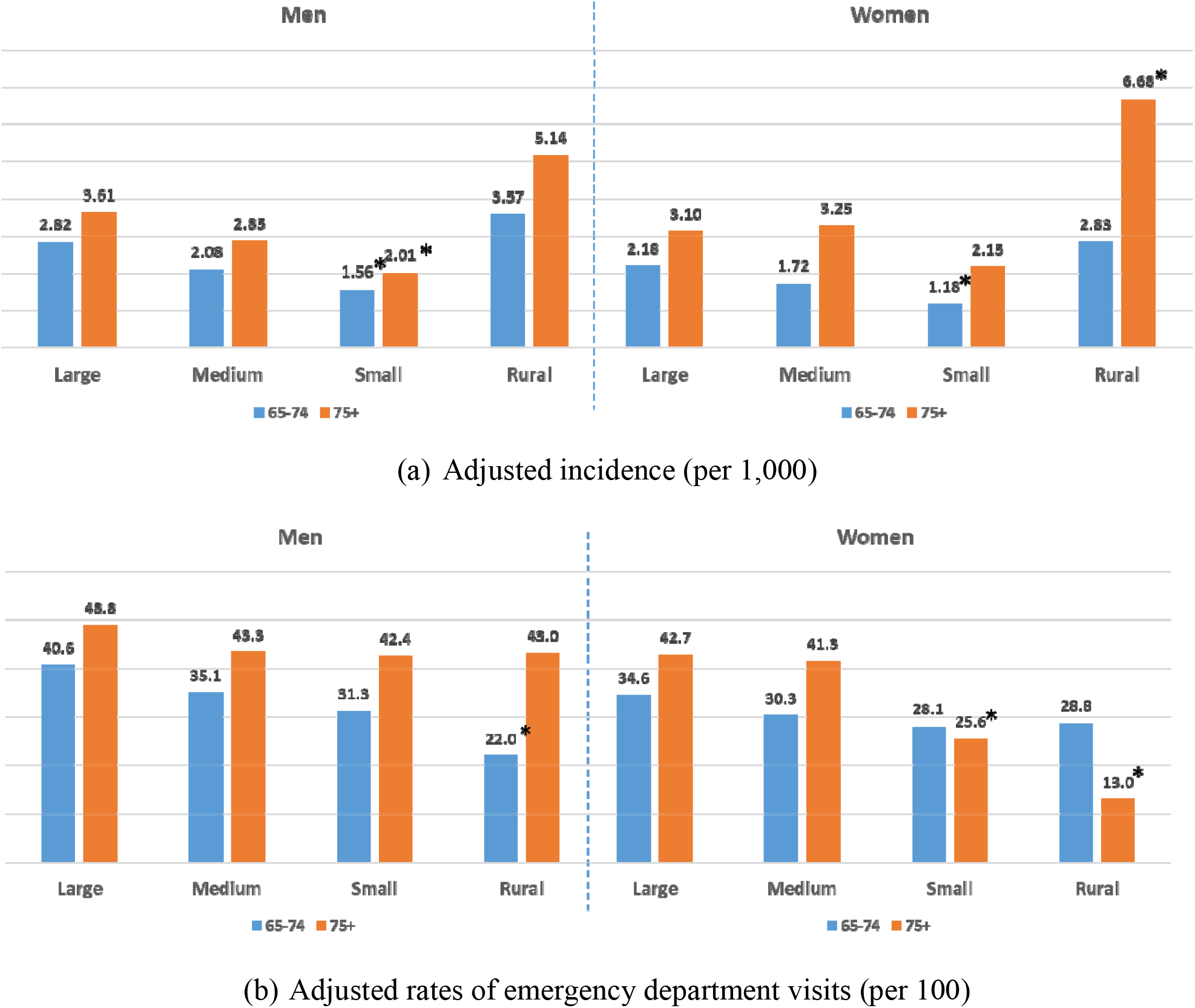
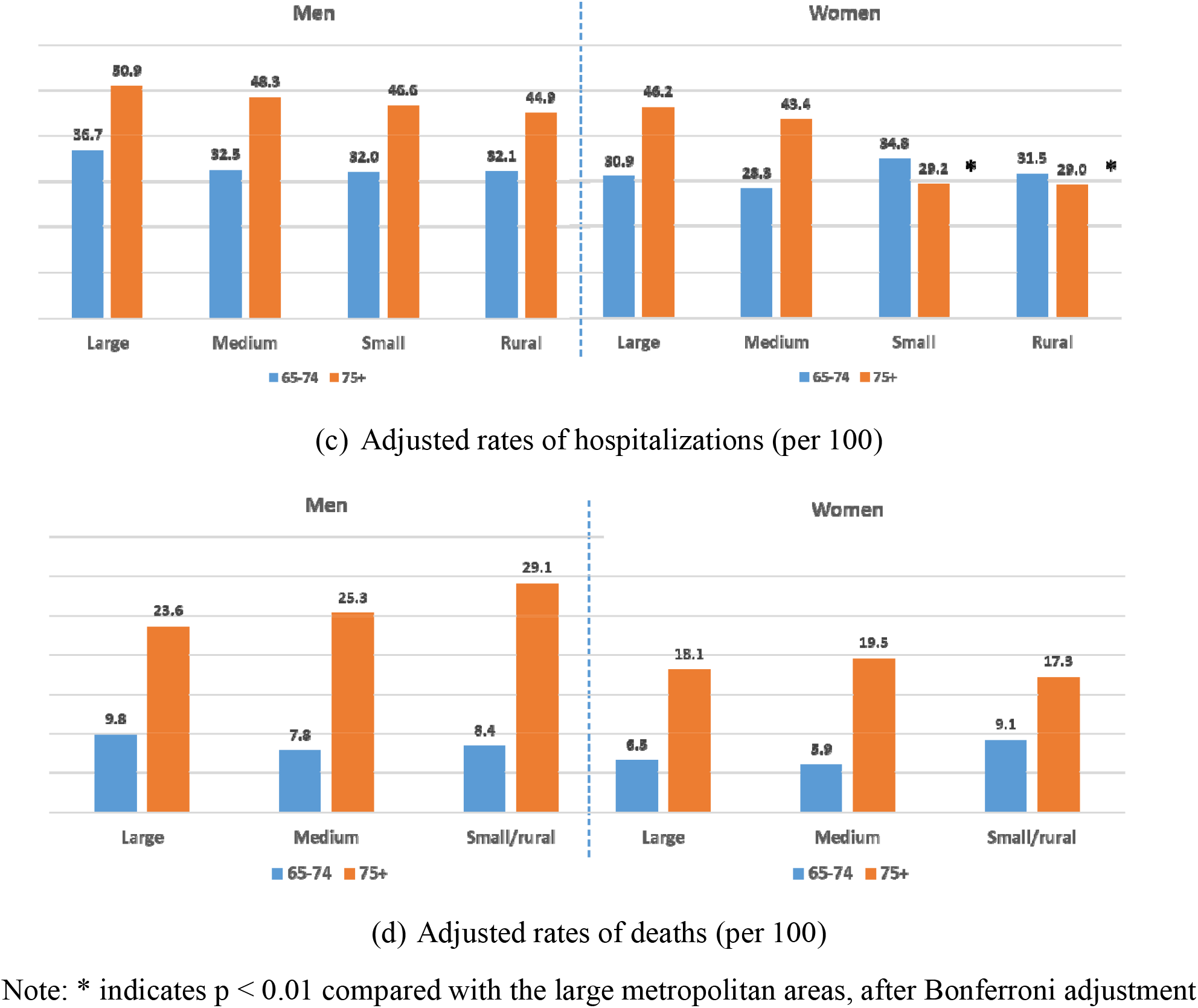
Adjusted incidence (per 1,000 persons) (a), and rates (per 100 cases) of emergency department visits (b), hospitalizations (c) and deaths (d) among elderly people with COVID-19, Florida as of May 27, 2020

Furthermore, those living in small metropolitan areas had lower incident rates than those living in large or medium metropolitan areas (Figure 2a, and Appendix Table 1). For example, for men aged 65-74 living in small metropolitan areas, the adjusted incidence was about 43% lower than that of large metropolitan areas (1.56 vs. 2.82 per 1,000 persons; Rate Ratio (RR): 0.57; 95% confidence interval (95%CI): [0.33-0.98]; p=0.04). Similar reduction was observed among women aged 65-74 (1.18 vs. 2.18 per 1,000 persons comparing small metropolitan with large metropolitan areas; RR: 0.53 [0.32-0.89]; p=0.02). However, elderly people living in rural areas tended to have higher incidence of COVID-19 than those living in large metropolitan areas. Particularly, women aged 75 or above living in rural areas had more than double of incidence than those living in large metropolitan areas (6.68 vs. 3.10 per 1,000 persons; RR: 2.23 [1.21-4.12]; p=0.01).

Figure 2b-d presented adjusted rates of ED visits, hospitalizations, and deaths by metropolitan statuses for each age and gender group. There were significant decreasing trends of ED visits across metropolitan statuses (all p for trend <0.01, Figure 2b, Appendix Table 2). For example, male patients aged 65-74 living in rural areas had 53% lower rate of ED visits than those living in large metropolitan areas (22% vs. 41%, OR: 0.47 [0.29-0.75]; p=0.002). Female patients aged 75 or above living in rural areas had 79% lower rate of ED visits than those living in large metropolitan areas (13% vs. 43%, OR: 0.21 [0.13-0.34], p<0.001). Similarly, those living in small metropolitan also had lower rates of ED visits than those living in large metropolitan areas.

Overall, there were decreasing trends of hospitalization rates across metropolitan statuses, but most evident among female patients aged 75 or above (Figure 2c, Appendix Table 2). Female patients aged 75 or above living in small metropolitan or rural areas had 53% and 63% lower rates of hospitalizations than those living in large metropolitan areas (29% vs. 46% for both comparisons, OR: 0.47 [0.34-0.66] and 0.37 [0.25-0.54] for small metropolitan and rural areas, respectively; both p <0.001).

Due to small number of deaths in small metropolitan and rural areas, we combined them in the analysis (Figure 2d, Appendix Table 2). There was no difference in death rates across metropolitan statuses.

The adjusted hospitalization rates were mapped by counties (Figure 3). In addition to Miami-Dade County (right southeast corner), there were pockets of small metropolitan or non-metro counties in the north or middle of Florida that had 40-60% or 60+% hospitalization rates (darker color).

**Figure 3:**
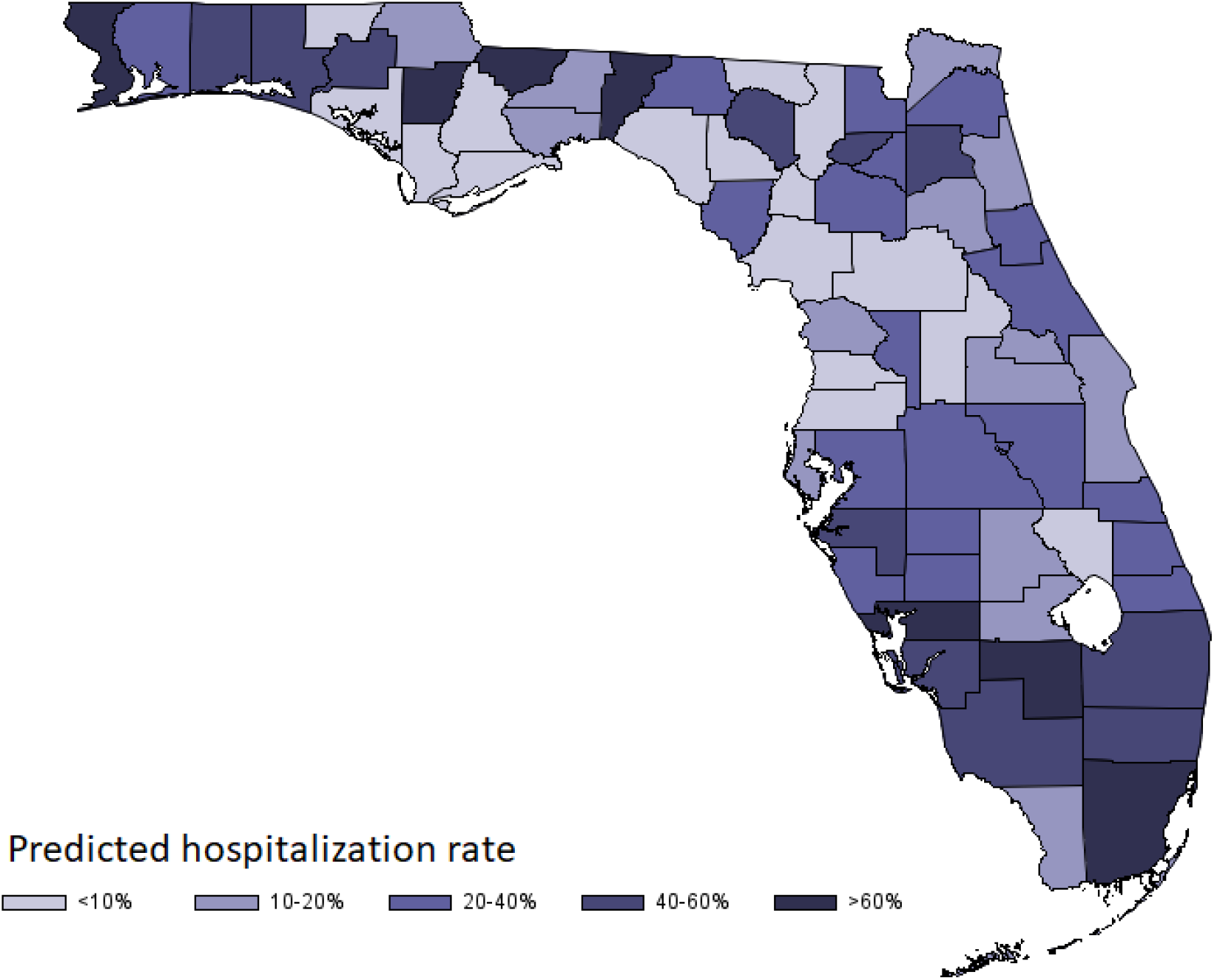
Choropleth map of hospitalization rates among people aged 65 or older by counties, Florida, as of May 27, 2020

## Discussion

This was the first study that documented significant health inequalities between large urban areas and small metropolitan or rural areas during the COVID-19 pandemic. In US Florida, elderly people living in small metropolitan areas had lower incidence of COVID-19, while those living in rural areas had higher incidence than those living in large or medium metropolitan areas. However, there were decreasing trends of ED visits and hospitalizations across metropolitan statuses. Particularly, the rates of ED visits and hospitalizations were significantly lower among female patients aged 75 or above living in small metropolitan or rural areas. On the other hand, the differences in COVID-19 related deaths were less evident between urban and non-urban areas. Many other factors could affect the risk of deaths due to COVID-19 and regional characteristics might play a less important role in mortality.

The reasons for lower incident COVID-19 rates in small metropolitan areas but higher incident rates in rural areas were complicated. The dispersed residence in small metropolitan and rural areas may deter the virus transmission, leading to relatively lower incident rates of COVID-19. On the other hand, there might not be enough detection kits available in these areas, resulting in artificially lower incident rates compared with large metropolitan areas. In addition, people living in rural areas might be more likely to rely on community centers and churches for social events. Regular gathering in community centers or churches might cause outbreaks of COVID-19, leading to abrupt increases of cases.

Furthermore, our findings confirmed deficiencies in providing health care to elderly people living outside of large or medium metropolitan areas (Casper et al. 2016; Singh et al. 2019). Health care facilities in the US were mostly concentrated in large cities. Elderly people living in small metropolitan or rural areas were known for lacking adequate health care (Odoi et al. 2019). In the time of emerging pandemic such as COVID-19, these problems may be aggravated when health care resources were under pressure. Many small metropolitan or rural hospitals were not equipped to manage infectious patients. Patients with mild symptoms might be triaged to self-care at home, without being diagnosed and lab confirmed. For elderly people, this was not ideal, as the respiratory symptoms might exacerbate suddenly (Huang et al. 2020a). Many of these severe cases were likely transferred to hospitals in larger cities, often enduring all kinds of troubles during the process.

Although the differences in the incident rates of COVID-19 between elderly men and women were small, there were gender differences in the rates of ED visits, hospitalizations and deaths among patients aged 75 or above living in small metropolitan or rural areas. This required careful explanations. It was unclear whether this was due to differences in disease severity, symptom tolerance, health care seeking behavior, or availability and accessibility to health care. Women were known to have lower tolerance of pain (Ruau et al. 2012), but this might not be applicable to infectious diseases.

Although our knowledge of COVID-19 was growing rapidly, the treatment outcomes were still unsatisfactory. Treating acute respiratory distress syndrome (ARDS) was still a major challenge, often leading to a mortality rate of 50% among those with ARDS (Huang et al. 2020a; Richardson et al. 2020). Many elderly patients, especially those with underlying conditions such as cardiovascular diseases and diabetes, often had more severe diseases than those who were young and healthy. Therefore, a coordinated public health system, together with timely virus detection, case isolation, symptom monitoring and active contact tracing, were more important to curb the epidemic. Small metropolitan and rural areas should not be overlooked in building this system.

This study had some limitations. First of all, not all patient’s information was publicly released due to privacy concerns. There were no explicit and accurate dates of symptom onset, clinic or ED visits, hospitalizations, and deaths for each patient. Therefore, we were only able to use logistic regressions to model the cumulative incidence of the ED visits, hospitalizations and deaths among those diagnosed with COVID-19. In addition, there was no information such as race and ethnicity, income and education levels in the file, hindering our ability to fully explore the roots of disparities (Hill et al. 2015) and precluding us from examine causalities of these health service inequalities. However, this problem was not unique to Florida. Many other states released aggregated data only. To some extent, we had more than enough data that were useful to paint a broad picture, but no good data to help us understand the drives of epidemic process and examine health disparities behind the case counts.

Second, this study was based on the existing data on the confirmed and reported cases, thus lacked people who were infected with virus but not reported (possibly asymptomatic or mild symptomatic). This may bias our results. Although elderly patients might be more likely to have symptoms if infected by the virus, we would nevertheless miss many asymptomatic or mildly symptomatic patients who would not seek care or not be detected. We did not know whether the proportion of asymptomatic patients differed between large metropolitan areas and small metropolitan or no-metro areas. In addition, the detection kits were not readily available to health providers, especially at the early stage of epidemic and in small metropolitan and rural areas. Therefore, patients living in small metropolitan or rural areas might be more likely to employ self-care or be triaged without diagnosis. We might underestimate the case incidences and over-estimate the rates of ED visits and hospitalizations among those living in small metropolitan or rural areas. The true health inequalities might be worse than our observed differences.

Third, this study only used Florida data because of the availability of individual cases. Although Florida has a larger percent of elderly population than many other US states, we should still be cautious to generalize our findings to other regions. In addition to the differences in population structure, other differences due to socio-economic status, physical environment (e.g., temperature, humidity, and altitude), and the scale of interventions and societal compliance to the interventions, may all influence health inequalities during the epidemic process.

Finally, the COVID-19 pandemic was still evolving. Although our previous research indicated that the epidemic had reached plateau since later April in the 30 US largest metropolitan areas (Yu 2020), there would still be a lot of new cases to come every day, as the instantaneous reproduction numbers remained around 1 in many US metropolitan areas right till May 27, 2020. The patterns of hospitalizations and deaths by different age, gender and regions would likely be more evident at the end of epidemic. Furthermore, given that much was still unknown regarding the treatments and consequences of COVID-19, elderly people might be impacted more profoundly by the epidemic and health inequalities due to possible long last disease consequences.

There were some unique strengths in our study. To our knowledge, this was the first study using individual patient information to examine urban and nonurban inequalities in the current COVID-19 epidemic. Health inequality issue was often neglected in the time of emerging epidemic, which were the reasons for recent urgent calls to tabulate cases and deaths by age, gender and ethnicities. Our research pointed to another dimension that should also be incorporated in epidemic reports. Furthermore, unlike common descriptive reports that focused on the numbers of new cases, hospitalizations and deaths, we employed analytical methods to uncover hidden health inequalities that were not evident in the aggregated tables. For example, comparing crude rates in Table 1 and adjusted rates in Appendix Table 2, only after careful adjustments did health inequalities emerge. Therefore, our study called for more good data, more transparent reporting, and more appropriate analyses.

In summary, profound health inequalities between urban and non-urban areas existed in the time of emerging pandemic like COVID-19. In US Florida, elderly people living in small metropolitan areas had lower incident rates, while elderly people living in rural areas had higher incident rates of COVID-19 than those living in large metropolitan areas. However, elderly patients living in small metropolitan or rural areas were less likely to have ED visits and hospitalizations than those living in large metropolitan areas, especially among female patients aged 75 or above living in these areas. Therefore, more supports and more resources should be granted to health care providers who serve the vulnerable populations in small metropolitan and rural areas.

## Data Availability

GitHub address later

## Ethics statement and declaration of conflict of interest

This study was deemed exempt from ethical review, as it used publicly available data and no human subjects were directly involved in the study. No inform consent was necessary. The authors declared no conflict of interest with respect to the research, authorship, and/or publication of this article.

## Acknowledgement

This study has benefited from many insightful comments from editors and several anonymous reviewers during the process of publication, leading to significant improvements in the final report.

## Appendix

**Appendix Figure 1:**
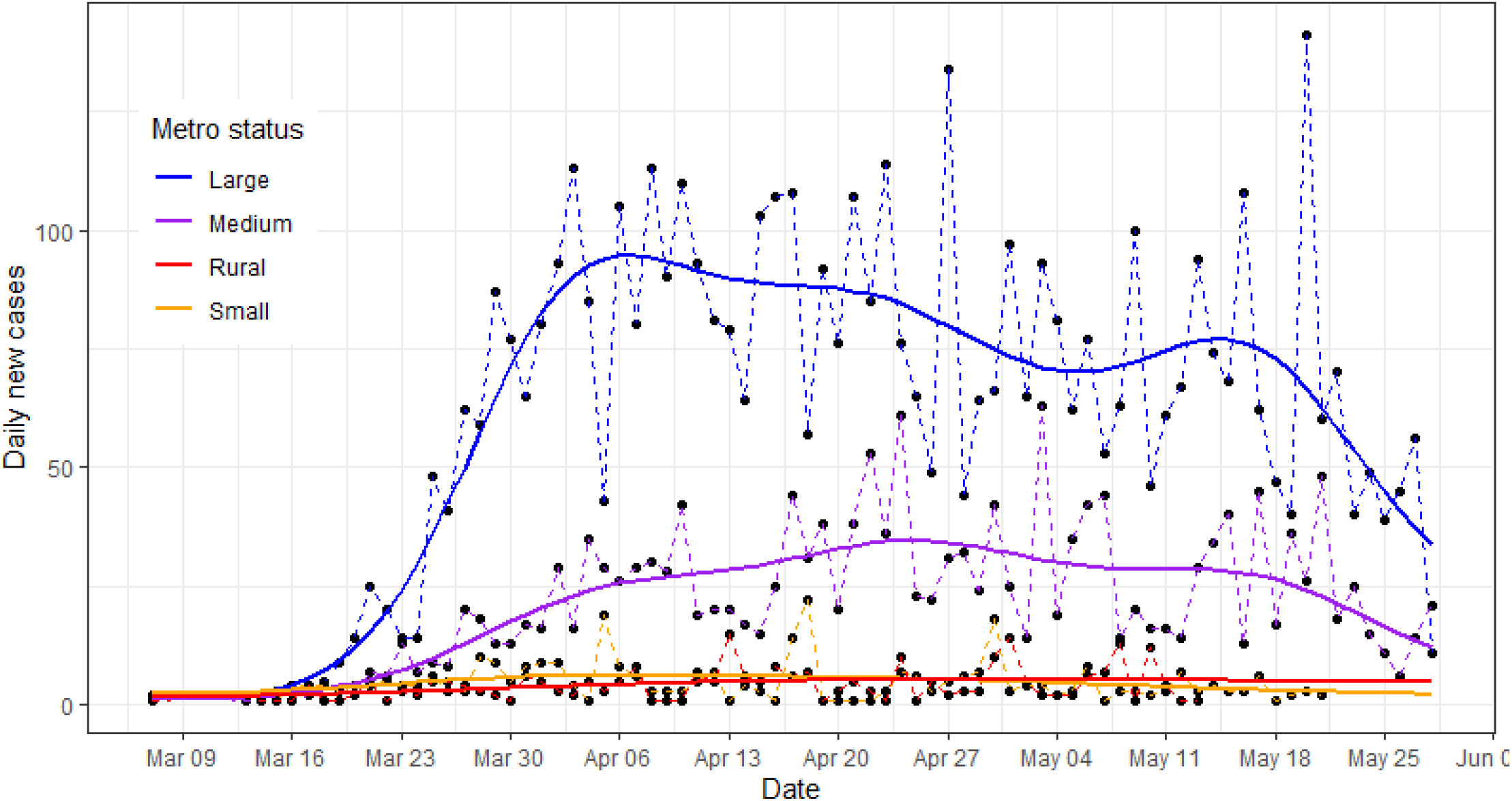
COVID-19 epidemic curves by metropolitan status among elderly people, FL

**Table 1:** COVID-19 confirmed cases, adjusted incidence (per 1,000) and 95% confidence interval (CI) among people aged 65 or older in Florida as of May 27, 2020

| **Gender & Age** | **Large metro cases** | **Large metro Incidence** | **Medium metro cases** | **Medium metro Incidence** | **Small metro cases** | **Small metro Incidence** | **Rural cases** | **Rural Incidence** | **p for trend** |
| --- | --- | --- | --- | --- | --- | --- | --- | --- | --- |
| **Total** | 9,492 | 2.93<br>(1.98 - 3.87) | 3,101 | 2.48<br>(1.73 - 3.23) | 615 | 1.73<br>(0.93 - 2.53) | 451 | 4.57<br>(3.19 - 5.95) | 0.10 |
| **Male** | 4,530 | 3.21<br>(2.17 - 4.25) | 1,437 | 2.46<br>(1.71 - 3.22) | 305 | 1.78*<br>(0.94 - 2.62) | 209 | 4.35<br>(2.97 - 5.74) | 0.29 |
| Age 65-74 | 2,316 | 2.82<br>(1.90 - 3.73) | 692 | 2.08<br>(1.44 - 2.73) | 151 | 1.56*<br>(0.81 - 2.31) | 108 | 3.57<br>(2.35 - 4.79) | 0.50 |
| Age 75 + | 2,214 | 3.61<br>(2.44 - 4.78) | 745 | 2.85<br>(1.97 - 3.73) | 154 | 2.01*<br>(1.04 - 2.98) | 101 | 5.14<br>(3.35 - 6.94) | 0.56 |
| **Female** | 4,962 | 2.64<br>(1.78 - 3.49) | 1,664 | 2.48<br>(1.73 - 3.24) | 310 | 1.67<br>(0.89 - 2.45) | 242 | 4.75*<br>(3.25 - 6.24) | 0.13 |
| Age 65-74 | 2,175 | 2.18<br>(1.47 - 2.88) | 647 | 1.72<br>(1.18 - 2.25) | 128 | 1.18*<br>(0.61 - 1.76) | 82 | 2.83<br>(1.80 - 3.85) | 0.65 |
| Age 75 + | 2,787 | 3.10<br>(2.09 - 4.11) | 1,017 | 3.25<br>(2.25 - 4.25) | 182 | 2.15<br>(1.13 - 3.18) | 160 | 6.68*<br>(4.49 - 8.87) | 0.02 |

Note: \* indicates $p < 0.05$ compared with the large metropolitan areas

**Table 2:**
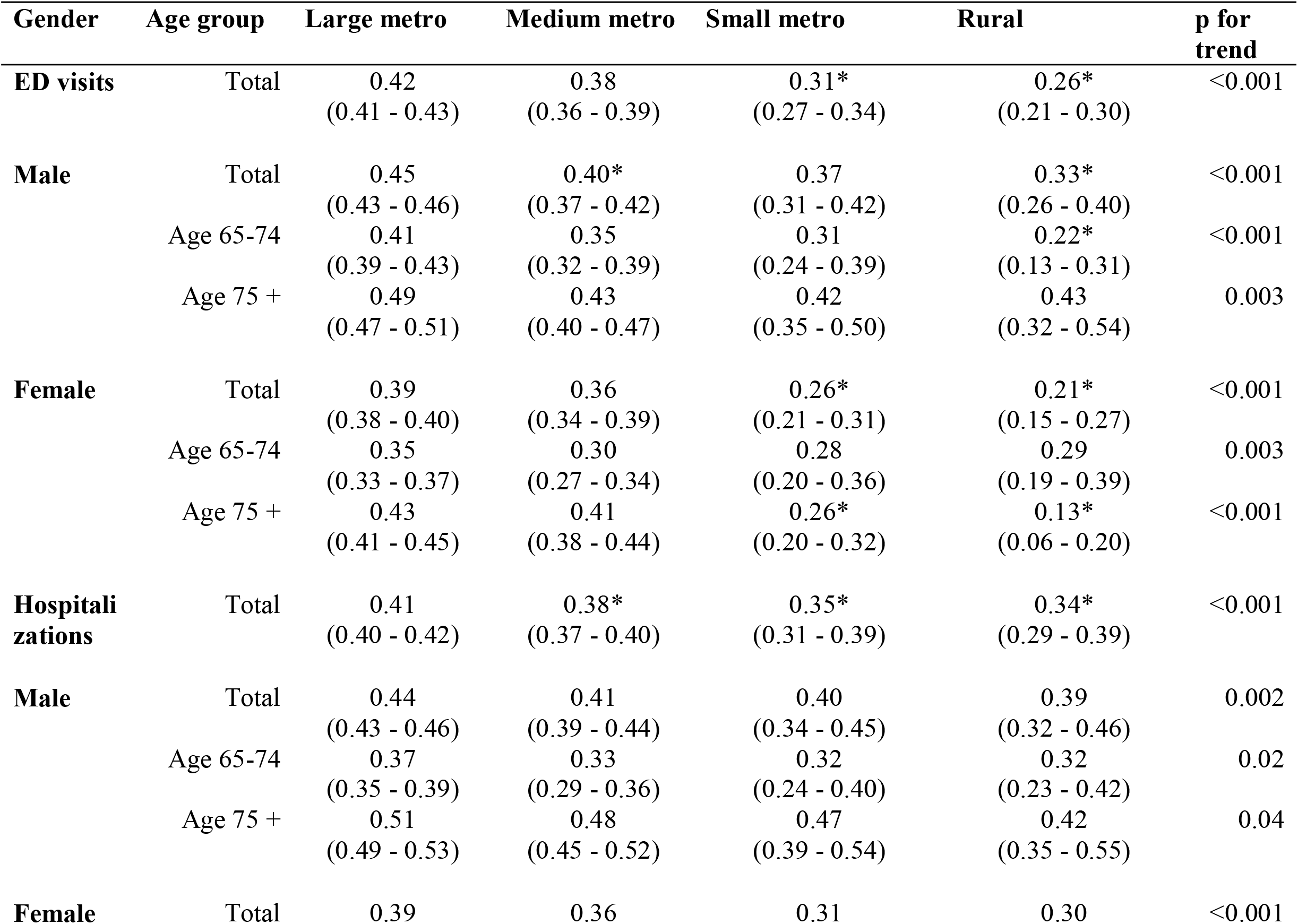
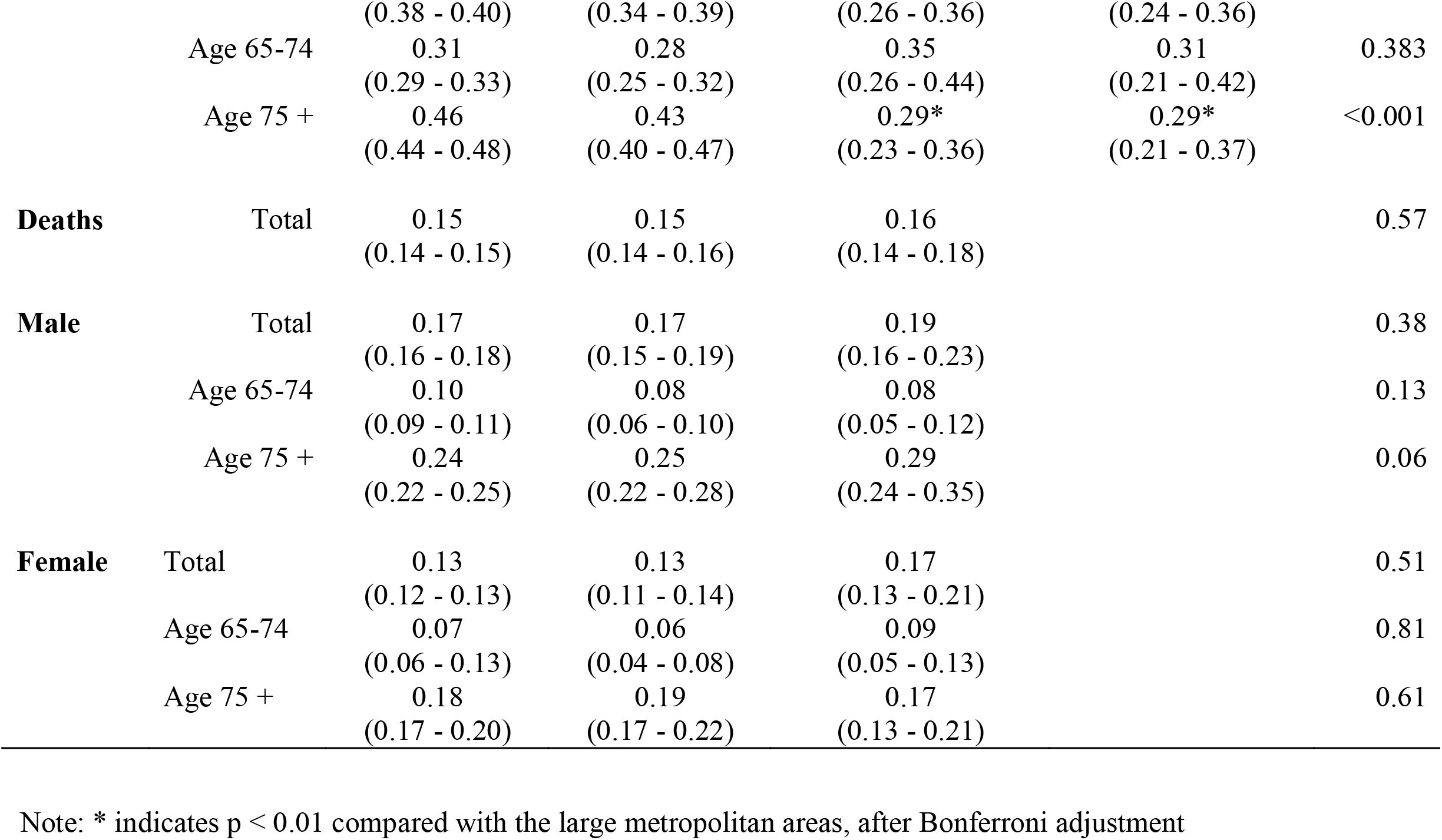
Predicted probabilities (rates) and 95% confidence interval (CI) of emergency department (ED) visits, hospitalizations and deaths among patients aged 65 or older in Florida, as of April 25, 2020

**Table 3:** Total number of people aged 65 or older infected with COVID-19 in Florida, as of May 27, 2020

| **County** | **Metropolitan status** | **COVID-19 Cases** |
| --- | --- | --- |
| Alachua | Medium metro | 91 |
| Baker | Large metro | 14 |
| Bay | Small metro | 16 |
| Bradford | Rural | 26 |
| Brevard | Medium metro | 113 |
| Broward | Large metro | 1576 |
| Calhoun | Rural | 37 |
| Charlotte | Small metro | 228 |
| Citrus | Small metro | 46 |
| Clay | Large metro | 148 |
| Collier | Medium metro | 351 |
| Columbia | Rural | 12 |
| Desoto | Rural | 32 |
| Dixie | Rural | 10 |
| Duval | Large metro | 345 |
| Escambia | Medium metro | 270 |
| Flagler | Medium metro | 51 |
| Franklin | Rural | . |
| Gadsden | Medium metro | 47 |
| Gilchrist | Medium metro | . |
| Glades | Rural | 6 |
| Gulf | Small metro | . |
| Hamilton | Rural | . |
| Hardee | Rural | 11 |
| Hendry | Rural | 60 |
| Hernando | Large metro | 27 |
| Highlands | Small metro | 34 |
| Hillsborou | Large metro | 487 |
| Holmes | Rural | . |
| Indian river | Small metro | 57 |
| Jackson | Rural | 12 |
| Jefferson | Medium metro | 15 |
| Lafayette | Rural | . |
| Lake | Large metro | 87 |
| Lee | Medium metro | 607 |
| Leon | Medium metro | 38 |
| Levy | Rural | 4 |
| Liberty | Rural | . |
| Madison | Rural | 9 |
| Manatee | Medium metro | 371 |
| Marion | Medium metro | 51 |
| Martin | Medium metro | 95 |
| Miami-Dade | Large metro | 4103 |
| Monroe | Rural | 24 |
| Nassau | Large metro | 18 |
| Okaloosa | Small metro | 68 |
| Okeechobee | Rural | 6 |
| Orange | Large metro | 264 |
| Osceola | Large metro | 154 |
| Palm Beach | Large metro | 1570 |
| Pasco | Large metro | 89 |
| Pinellas | Large metro | 433 |
| Polk | Medium metro | 305 |
| Putnam | Rural | 31 |
| St. Johns | Large metro | 60 |
| St. Lucie | Medium metro | 126 |
| Santa Rosa | Medium metro | 49 |
| Sarasota | Medium metro | 303 |
| Seminole | Large metro | 114 |
| Sumter | Small metro | 131 |
| Suwannee | Rural | 69 |
| Taylor | Rural | . |
| Union | Rural | 7 |
| Volusia | Medium metro | 211 |
| Wakulla | Medium metro | 3 |
| Walton | Small metro | 35 |
| Washington | Rural | 26 |

